# Impact of rotavirus vaccination in Malawi from 2012 to 2022 compared to model predictions before, during, and after the COVID-19 pandemic

**DOI:** 10.1101/2024.05.29.24308124

**Authors:** Virginia E. Pitzer, Latif Ndeketa, Ernest O. Asare, Daniel Hungerford, Khuzwayo C. Jere, Nigel A. Cunliffe

## Abstract

**Background:** Rotarix® rotavirus vaccine was introduced into the Malawi national immunization program in October 2012. We used a previously developed mathematical models to estimate overall vaccine effectiveness over a 10-year period following rotavirus vaccine introduction.

**Methods:** We analyzed data on children <5 years old hospitalized with acute gastroenteritis in Blantyre, Malawi from January 2012 to June 2022, compared to pre-vaccination data. We estimated vaccine coverage before, during, and after the COVID-19 pandemic using data from rotavirus-negative children. We compared model predictions for the weekly number of rotavirus-associated gastroenteritis (RVGE) cases to the observed number by age to validate model predictions and estimate overall vaccine effectiveness.

**Results:** The number of RVGE and rotavirus-negative acute gastroenteritis cases declined substantially following vaccine introduction. Vaccine coverage among rotavirus-negative controls was >90% with two doses by July 2014, and declined to a low of ∼80% in October 2020, before returning to pre-pandemic levels by July 2021. Our models captured the post-vaccination trends in RVGE incidence, with 5.4% to 19.4% of observed weekly RVGE cases falling outside of the 95% prediction intervals. Comparing observed RVGE cases to the model-predicted incidence without vaccination, overall vaccine effectiveness was estimated to be 36.0% (95% prediction interval: 33.6%, 39.9%) peaking in 2014 and was highest in infants (52.5%; 95% prediction interval: 50.1%, 54.9%).

**Conclusions:** Overall effectiveness of rotavirus vaccination in Malawi is modest despite high vaccine coverage and has plateaued since 2016. Our mathematical models provide a validated platform for assessing strategies to improve rotavirus vaccine impact.

## Background

Rotavirus is the leading cause of severe diarrhea among children around the world [1,2]. While rotavirus vaccines have been introduced into over half of all countries worldwide [3], vaccine-induced immunity is non-sterilizing and vaccine efficacy and effectiveness tend to be lower in low-income settings [4,5]. As a result, rotavirus is still the most common cause of acute watery diarrhea among infants even in countries with a rotavirus vaccination program [6].

In Malawi, the monovalent Rotarix® vaccine (RV1; GlaxoSmithKline) was introduced into the national immunization program on October 29, 2012, with two doses administered orally at 6 and 10 weeks of age [7]. In the Malawi cohort of a phase III clinical trial, vaccine efficacy was 38.1% over two years of follow-up [8]. Estimates of vaccine effectiveness have been slightly higher (61.9%-70.6%), but the overall reduction in rotavirus prevalence among children hospitalized with diarrhea in Blantyre, Malawi has been modest (from 32% pre-vaccine to 24-28% post-vaccine) despite high vaccine coverage (>84% over since 2013) [9–11]. Using a mathematical model fitted to pre- and post-vaccination data from Blantyre, we previously showed that the observed vaccine effectiveness and impact of vaccination could be explained by heterogeneity in vaccine response possibly combined with waning of vaccine-induced immunity [12].

The COVID-19 pandemic led to disruptions in vaccine delivery and healthcare seeking behavior throughout the world [13]. Across the African Region, coverage with first dose of diphtheria-tetanus-pertussis containing vaccine (DTP1) was estimated to decline by 3% (from 83% in 2019 to 80% in 2021-2022), while coverage with the third dose (DTP3) declined by 5% (from 77% to 72%) [14]. In Malawi, one study reported a 20.0% decline in inpatient care and a 17.6% decline in immunization services between April 2020 and December 2021 relative to the pre-pandemic period (2015-2019) [15], whereas other studies have reported little to no disruption in maternal health services and cumulative vaccine doses administered [16,17].

Here, we examine trends in rotavirus-associated gastroenteritis (RVGE) cases by age group over the 10-year period after vaccine introduction in Blantyre, Malawi. Using data from children with rotavirus-negative acute gastroenteritis (test-negative controls), we estimate RV1 coverage before, during, and after the COVID-19 pandemic. By comparing the observed number of RVGE cases by age group to predictions from our previously developed mathematical model, we validate model predictions based on five years of out-of-sample data from 2017 to 2022. Finally, we estimate vaccine impact by comparing the observed RVGE incidence to that predicted by the mathematical model in the absence of vaccination.

## Methods

Surveillance for RVGE has been conducted in Blantyre, Malawi among inpatients and outpatients at Queen Elizabeth Central Hospital (QECH) from July 1997 to June 2007, January 2008 to December 2009 [18,19], and since January 2012 [10,20], as previously described. Briefly, children <5 years of age who presented to QECH with acute gastroenteritis (AGE, defined as three or more loose stools in a 24-hour period for less than 14 days) and moderate to severe dehydration were enrolled, fecal samples were collected, and rotavirus testing was carried out using a sensitive and specific enzyme-linked immunosorbent assay (ELISA; Rotaclone®; Meridian Bioscience); the G- and P-types of rotavirus-positive samples were determined using reverse-transcriptase polymerase chain reaction (RT-PCR) [21]. We analyzed aggregate data on the number of rotavirus-positive and rotavirus-negative AGE cases in each week of the surveillance period through June 2022 for 1-month age categories <2 years of age and 1-year age categories for 2 to <5 years of age.

Sample collection was halted between April and early October 2020 due to the COVID-19 pandemic. Following the pandemic, healthcare seeking for diarrhea at QECH substantially declined before gradually increasing to pre-pandemic levels in 2022. To control for changes in healthcare seeking and reporting effort over time, we calculated the 2-year (105-week) moving average of the number of rotavirus-negative AGE cases in the <1 year, 1 to <2 year, and 2 to <5 years age groups; we then divided by the mean number of rotavirus-negative AGE cases for the entire surveillance period in each age group to estimate the relative reporting rate over time.

Vaccination status was recorded on government-issued family-held records. To estimate vaccine coverage over time, we divided the number of age-eligible rotavirus-negative AGE cases who had received one or two doses of the RV1 vaccine by the total number of age-eligible rotavirus-negative AGE cases who presented to QECH in the 9-month (39-week) period centered on each week from October 28, 2012 (week of vaccine introduction) to June 2022. Uncertainty was quantified using 95% binomial confidence intervals.

To estimate the counterfactual number of RVGE cases that would be expected in each age group over time had the RV1 vaccine not been introduced, we used mathematical models that we previously developed and fitted to pre- and post-vaccination RVGE cases at QECH (Figure S1, Figure S2). Details of the modeling approach have been previously described [12]. Briefly, in the absence of vaccination, the model assumes that infants are born with maternal immunity, which wanes after a period of ∼6 months. After each rotavirus infection, individuals are briefly immune to reinfection, then gain partial acquired immunity that reduces the severity and rate of subsequent infections; we assume the risk of moderate-to-severe RVGE is negligible following two natural infections. The model was parameterized based on data from birth cohort studies (see Table S1), then fitted to the 12 years of pre-vaccination data from QECH to estimate the mean transmission rate (as quantified by the basic reproductive number, *R*_0_), amplitude and timing of seasonality in transmission, and the proportion of moderate-to-severe RVGE cases in Blantyre that seek care at QECH and are tested (i.e. reporting fraction). To control for variation in reporting effort over time, we multiplied the reporting fraction by the relative reporting rate (described above). We initially fit the model via maximum a posterior estimation, then used a Markov chain Monte Carlo to obtain samples from the posterior distributions of model parameters (Table S1) [12].

To model the effect of vaccination with RV1, we explored different assumptions about the probability of responding to each vaccine dose and the potential waning of vaccine-induced immunity. Models 1 and 2 assume that each vaccine dose provides immunity comparable to one natural infection among those who respond to the vaccine, i.e. partial immunity against reinfection and full protection against moderate-to-severe RVGE following two “successful” vaccine doses and/or natural infections. Model 1 assumes the proportions of infants who respond to the first and second dose are independent of one another (i.e. homogeneity in vaccine response), whereas Model 2 assumes that individuals who fail to respond to the first dose may be less likely to respond to subsequent doses (i.e. heterogeneity in vaccine response). The probability of responding to each dose was estimated based on seroconversion data from a vaccine trial conducted in Malawi [8]; uncertainty was characterized using beta distributions, as detailed previously (Table S2) [12]. Neither model was fitted to the post-vaccination data.

Models 3 and 4 assume each vaccine dose provides, among those who respond, temporary but complete immunity against rotavirus infection. Following the waning of vaccine-induced immunity, vaccinated infants return to their previous level of susceptibility, while those who respond to both vaccine doses move to the next vaccinated (and protected) compartment.

Model 3 assumes homogeneity in vaccine response, whereas Model 4 assumes heterogeneity in vaccine response. The probability of responding to each vaccine dose and the rate of waning of vaccine-induced immunity were estimated by fitting to the post-vaccination data from QECH between October 2012 and August 2017 (Table S2) [12].

To validate model predictions for the impact of rotavirus vaccination in Malawi, we compared the predicted number of RVGE cases from the four models to the observed number of rotavirus-positive AGE cases at QECH. We considered both the total number of cases per week among children <5 years old as well as cases in each of the three age groups (<1 year, 1 to <2 years, and 2 to <5 years of age). First, we calculated the Spearman rank correlation coefficient and root mean square error (RMSE) between the model-predicted mean number of RVGE cases stratified by week and age group and the observed number of RVGE cases per week and age group for three time periods: the full 10-year time period from January 2012 to June 2022 (excluding the period from April-October 2020 with no surveillance); the in-sample validation period (January 2012-August 2017, previously used for fitting Models 3 and 4); and the out-of-sample validation period (September 2017-June 2022). Second, we calculated the proportion of weeks in which the 95% prediction interval for the model-predicted number of RVGE cases contained the observed number of RVGE cases. To generate the model predictions, we sampled 100 times from the joint posterior distributions of the pre- and post-vaccination model parameters. To account for observation error around the mean number of RVGE cases predicted for each week (comparable to the approach used for model fitting), we sampled 100 times from a Poisson distribution with rate parameter *λ_w,a_* equal to the mean number of RVGE cases in week *w* and age group *a* for each of the 100 model simulations. The 95% prediction intervals were generated based on the 2.5 and 97.5 percentiles of the resulting 10,000 samples for each week (and within each age group) for the different models.

Using the validated models, we then estimated the overall vaccine effectiveness for each year following the introduction of RV1 in 2012. For each of the models, we simulated both the predicted mean number of RVGE cases given the estimated vaccine coverage over time as well as the mean number of RVGE cases predicted with no vaccination (i.e. assuming vaccine coverage was 0%). All four models predicted the same number of RVGE cases in the absence of vaccination, as expected. Overall vaccine effectiveness (OE), defined as the reduction in disease incidence among both vaccinated and unvaccinated individuals in a vaccinated population compared to an unvaccinated population [22], in year *y* and age group *a* was estimated as:

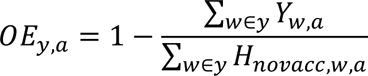

where *Y_w,a_*is the observed number of RVGE cases in week *w* (in year *y*) and age group *a* and *H_novacc,w,a_* is the model-predicted number of RVGE cases in week *w* and age group *a* with no vaccination.

Code for the model and analysis was written and implemented in MATLAB v9.14 (Mathworks, Natick, MA) and is available at https://github.com/vepitzer/rotavirusMalawi.

## Results

Since the introduction of rotavirus vaccination in October 2012, the number of RVGE cases per year at QECH has declined from 193 in 2013 to 36 in 2021 (Table 1, Figure S3). However, the number of rotavirus-negative AGE cases also declined over this time period, from 530 in 2013 to 71 in 2021. Thus, not all of the decline in the incidence of RVGE cases can be attributed to the RV1 vaccine. There has been reduced healthcare seeking for diarrhea at QECH, particularly since the onset of the COVID-19 pandemic in 2020 (Table 1, Figures S3-S4).

**Table 1.** Number of rotavirus-positive and rotavirus-negative acute gastroenteritis cases diagnosed at Queen Elizabeth Central Hospital in Blantyre, Malawi, 2012-2022. Values in parentheses are the percent of cases in each age group.

| Year | Rotavirus-positive |  |  |  | Rotavirus-negative |  |  |  |
| --- | --- | --- | --- | --- | --- | --- | --- | --- |
|  | <5y | <1y | 1-<2y | 2-<5y | <5y | <1y | 1-<2y | 2-<5y |
| Pre-vaccination (1997-2009)* | 39-235 | 18-189<br>(79.4%) | 8-46<br>(18.6%) | 0-5<br>(2.0%) | 116-423 | 44-258<br>(60.8%) | 29-121<br>(29.5%) | 11-44<br>(9.7%) |
| 2012 | 192 | 149<br>(77.6%) | 40<br>(20.8%) | 3<br>(1.6%) | 315 | 170<br>(54.0%) | 84<br>(26.7%) | 61<br>(19.4%) |
| 2013 | 193 | 120<br>(62.2%) | 65<br>(33.7%) | 8<br>(4.2%) | 530 | 328<br>(61.9%) | 162<br>(30.6%) | 40<br>(7.6%) |
| 2014 | 137 | 81<br>(59.1%) | 52<br>(38.0%) | 4<br>(2.9%) | 648 | 318<br>(49.1%) | 260<br>(40.1%) | 70<br>(10.8%) |
| 2015 | 161 | 90<br>(55.9%) | 60<br>(37.3%) | 11<br>(6.8%) | 428 | 225<br>(52.6%) | 141<br>(32.9%) | 62<br>(14.5%) |
| 2016 | 128 | 80<br>(62.5%) | 41<br>(32.0%) | 7<br>(5.5%) | 235 | 143<br>(60.9%) | 68<br>(28.9%) | 24<br>(10.2%) |
| 2017 | 111 | 72<br>(64.9%) | 29<br>(26.1%) | 10<br>(9.0%) | 235 | 129<br>(54.9%) | 83<br>(35.3%) | 23<br>(9.8%) |
| 2018 | 110 | 62<br>(56.4%) | 42<br>(38.2%) | 6<br>(5.5%) | 225 | 124<br>(55.1%) | 83<br>(36.9%) | 18<br>(8.0%) |
| 2019 | 76 | 46<br>(60.5%) | 26<br>(34.1%) | 4<br>(5.3%) | 154 | 91<br>(59.1%) | 50<br>(32.5%) | 13<br>(8.4%) |
| 2020<sup>#</sup> | 26 | 11<br>(42.3%) | 13<br>(50.0%) | 2<br>(7.7%) | 64 | 38<br>(59.4%) | 19<br>(29.7%) | 7<br>(10.9%) |
| 2021 | 36 | 21<br>(58.3%) | 12<br>(33.3%) | 3<br>(8.3%) | 71 | 40<br>(56.3%) | 21<br>(29.6%) | 10<br>(14.1%) |
| 2022<sup>§</sup> | 26 | 16<br>(61.5%) | 10<br>(38.5%) | 0<br>(0%) | 46 | 24<br>(52.2%) | 16<br>(34.8%) | 6<br>(13.0%) |
| Post-vaccination<br>(2013-2022<sup>§</sup>) | 1004 | 599<br>(59.7%) | 350<br>(34.9%) | 55<br>(5.5%) | 2636 | 1460<br>(55.4%) | 903<br>(34.3%) | 273<br>(10.4%) |

\* The range of yearly values and average age distribution is presented for the pre-vaccination period.
<sup>#</sup> Excludes the period from April 5 to October 10 when surveillance was halted because of the COVID-19 pandemic.
<sup>§</sup> Limited to data through June 25, 2022.

There has also been a shift in the age distribution of RVGE cases. Prior to vaccine introduction, nearly 80% of RVGE cases occurred among infants <1 year old, with 18.6% occurring among 1-year-olds and only 2% among 2- to <5-year-olds (Table 1). Since 2013, the proportion of RVGE cases among <1-year-olds has consistently been <65%, with a third of cases occurring among 1-year-olds and 5% occurring among 2- to <5-year-olds. While there has also been a slight shift in the burden of non-rotavirus AGE cases to older age groups, it has been less marked (Table 1).

Vaccine coverage among non-rotavirus AGE controls at QECH reached high levels by January 2015 (approximately two years after vaccine introduction), with >95% having received at least one dose of RV1 and >90% having received two doses. However, there was a slight decline in vaccine coverage during the COVID-19 pandemic in 2020-2021, particularly for the second dose, although coverage has since rebounded to pre-pandemic levels (Figure 1).

**Figure 1.**
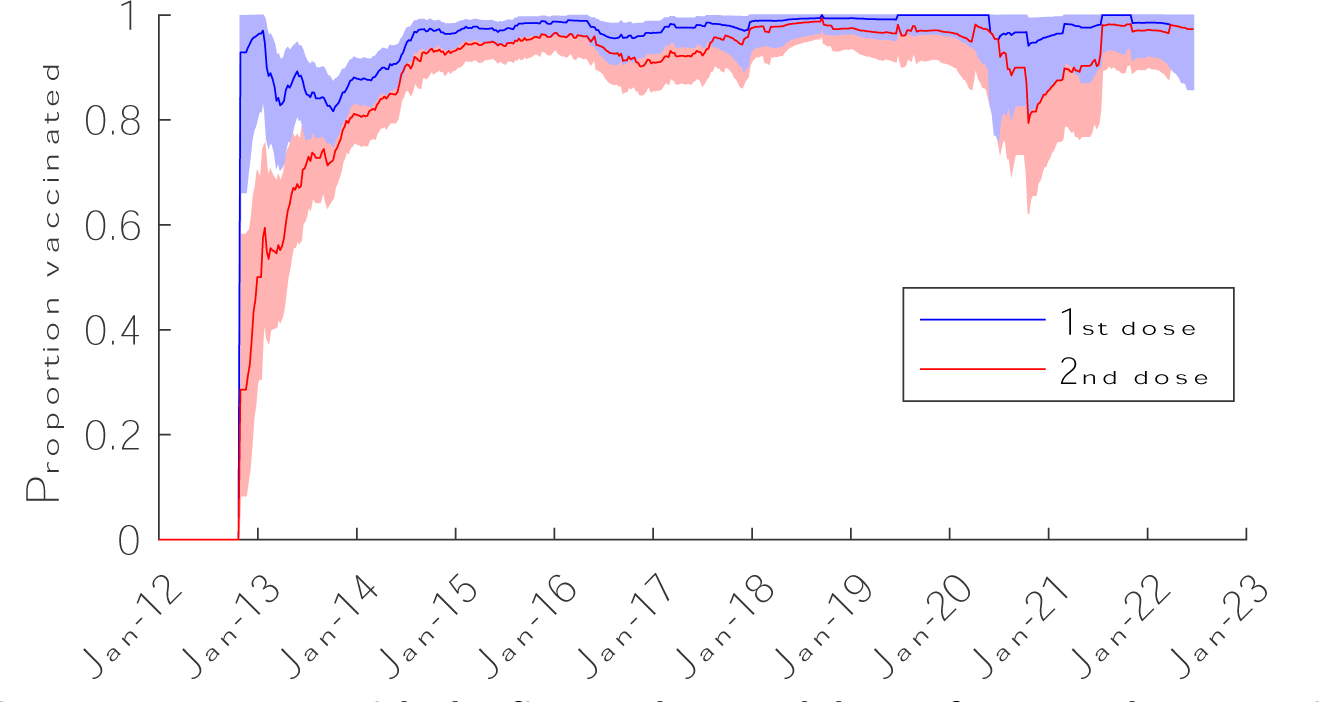
Coverage with the first and second dose of monovalent rotavirus vaccine among test-negative controls at Queen Elizabeth Central Hospital, January 2012-June 2022. The 39-week moving average of the proportion of age-eligible rotavirus-negative acute gastroenteritis cases who had received one dose (blue) and two doses (red) of the RV1 vaccine are plotted. The shaded regions represent the 95% binomial confidence intervals.

Our mathematical models were able to capture the post-vaccination trends in RVGE incidence, although the model assuming homogeneity in vaccine response and no waning of vaccine-induced immunity (Model 1) tended to overestimate vaccine impact and performed less well (Figure 2, Figures S5). Observed and model-predicted RVGE cases were moderately correlated (Spearman’s *ρ* = 0.54-0.56 for Models 1-4), and the RMSE was similar across the four models, with Model 2 exhibiting the lowest RMSE (Table 2). The percent of weeks in which the observed number of RVGE cases fell outside the 95% prediction interval of the models was approximately 5% for Models 3 and 4 (as expected). The percentage was slightly higher for Models 1 and 2, particularly for the 1- to <2-year-old and 2- to <5-year-old age groups (Table 2); however, it is worth noting that these models were not fitted to the post-vaccination data, whereas Models 3 and 4 were fitted to data through August 2017. The percent of weeks in which the observed number of RVGE cases fell outside the 95% prediction intervals was similar for the period before and after August 2017 (Table 2).

**Figure 2.**
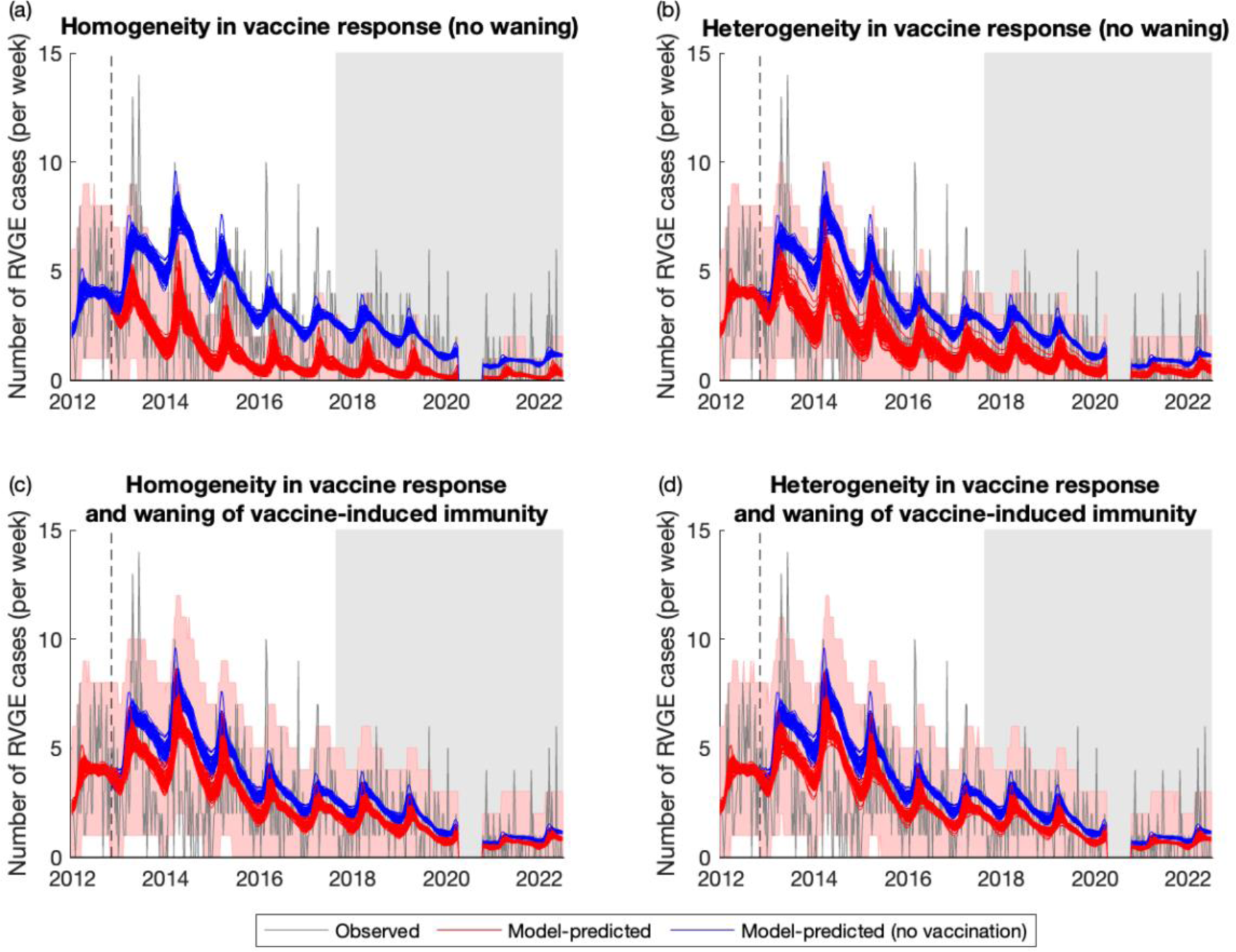
Weekly timeseries of observed and model-predicted rotavirus gastroenteritis cases at Queen Elizabeth Central Hospital, January 2012-June 2022. The observed number of RVGE cases per week is plotted in grey, while model predictions for the average weekly number of RVGE cases given current estimates of vaccine coverage (red lines) and assuming no vaccination (blue lines) are plotted for 100 samples from the posterior distribution of model parameters for (a) Model 1, (b) Model 2, (c) Model 3, and (d) Model 4. The red shaded region represents the 95% prediction intervals assuming the observed number of cases per week are Poisson distributed. The dashed vertical line shows the week of vaccine introduction, while the light grey shaded region shows the out-of-sample validation period.

**Table 2.** Validation of model predictions for the impact of vaccination on RVGE cases for January 2012-June 2022 and the in-sample (January 2012-August 2017) and out-of-sample (August 2017-June 2022) periods.

|  | Time period | Spearman's correlation between observed & predicted RVGE cases | Root mean square error | Percent of weeks in which the observed number of RVGE cases fell outside the 95% prediction interval |  |  |  |
| --- | --- | --- | --- | --- | --- | --- | --- |
|  |  |  |  | All ages | <1 year old | 1-<2 years old | 2-<5 years old |
| **Model 1:** no waning, homogeneous response | Entire period | 0.541 | 1.108 | 19.4% | 8.7% | 12.3% | 7.5% |
|  | In-sample | 0.561 | 1.293 | 19.3% | 8.8% | 12.2% | 7.8% |
|  | Out-of-sample | 0.402 | 0.804 | 19.6% | 8.4% | 12.4% | 7.1% |
| **Model 2:** no waning, heterogeneous response | Entire period | 0.560 | 1.058 | 8.1% | 2.7% | 7.5% | 7.1% |
|  | In-sample | 0.579 | 1.246 | 8.5% | 2.7% | 6.4% | 7.1% |
|  | Out-of-sample | 0.417 | 0.743 | 7.6% | 2.7% | 8.9% | 7.1% |
| **Model 3:** waning vaccine immunity, homogeneous response | Entire period | 0.562 | 1.072 | 5.4% | 2.5% | 4.0% | 0.6% |
|  | In-sample | 0.581 | 1.280 | 6.8% | 3.4% | 3.1% | 1.0% |
|  | Out-of-sample | 0.427 | 0.711 | 3.6% | 1.3% | 5.3% | 0.0% |
| **Model 4:** waning vaccine immunity, heterogeneous response | Entire period | 0.562 | 1.077 | 5.4% | 2.3% | 4.6% | 0.4% |
|  | In-sample | 0.584 | 1.287 | 6.8% | 3.1% | 3.7% | 1.0% |
|  | Out-of-sample | 0.424 | 0.715 | 3.6% | 1.3% | 6.2% | 0.0% |

Compared to the model-predicted number of RVGE cases with no vaccination, the observed number of RVGE cases among all children <5 years of age following the introduction of RV1 was reduced by 36.0% (95% prediction interval: 33.6%, 39.9%). The overall vaccine effectiveness (OE) was greatest in the <1-year-olds (52.5%; 95% prediction interval: 50.1%, 54.9%), whereas the post-vaccination incidence of RVGE was slightly higher than predicted under the no-vaccination scenario for 1-<2-year-olds (OE=-18.6%; 95% prediction interval: −33.4%, −5.6%) and for 2-<5-year-olds (OE=-336.4%; 95% prediction interval: −238.8%, −471.9%). Overall effectiveness was greatest in 2014 (two years after vaccine introduction) and lowest in 2020, driven primarily by an increase in cases among 1-<2-year-olds (Figure 3). Since 2017, overall vaccine effectiveness has remained fairly consistent (Table S3), with an average overall effectiveness of 14.5% (95% prediction interval: 10.2%, 19.7%) and an average overall effectiveness in <1-year-olds of 36.4% (95% prediction interval: 33.0%, 39.2%).

**Figure 3.**
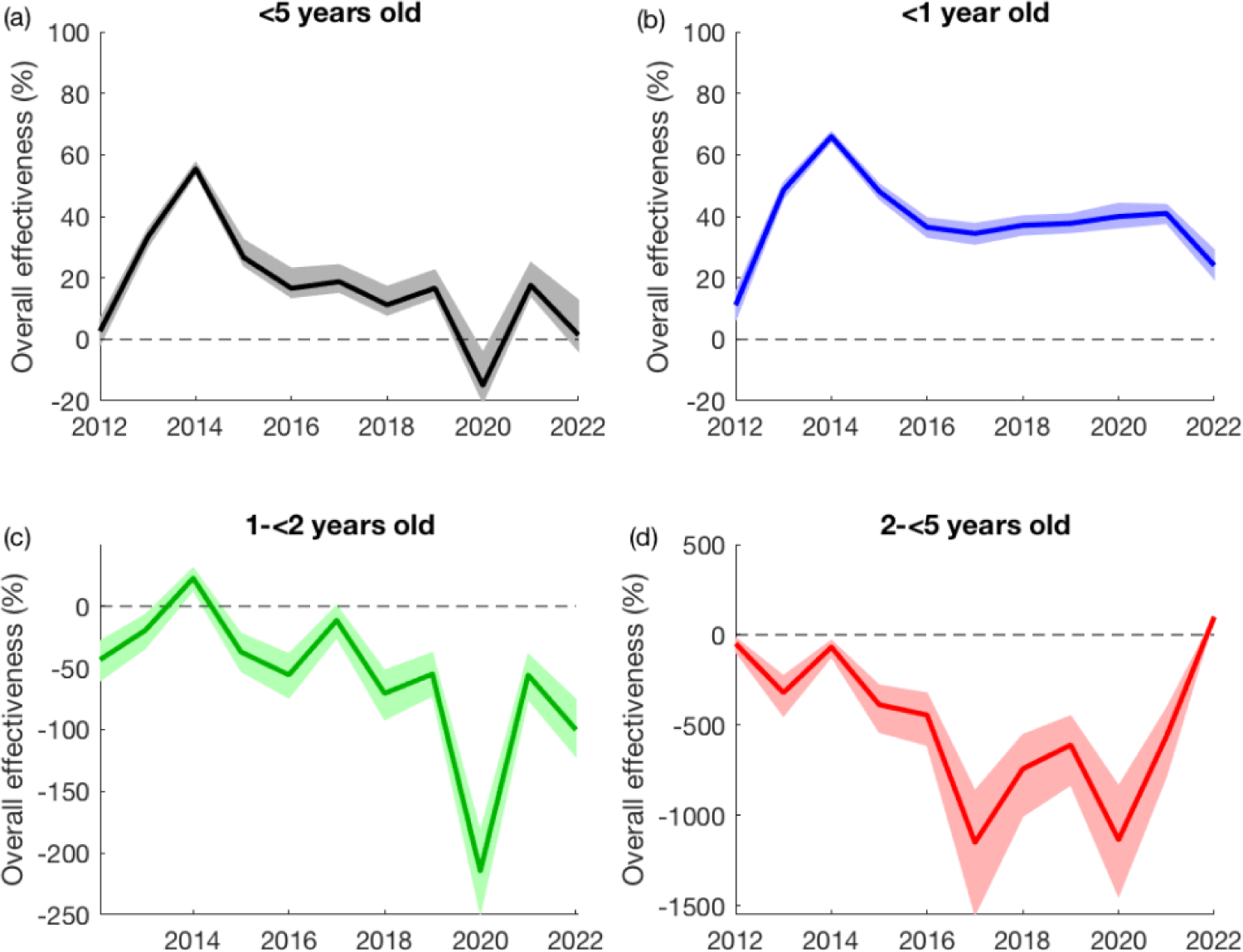
Model-based estimates of overall vaccine effectiveness by year and age group. The overall effectiveness is estimated by comparing the observed number of RVGE cases in each year to the model-predicted number of RVGE cases per year assuming no vaccination and is plotted for all children <5 years old (a) and by age group: (b) <1 year old in blue, (c) 1 to <2 years old in green, and (d) 2 to <5 years old in red. The shaded regions represent the 95% prediction intervals.

## Discussion

It has been over 10 years since rotavirus vaccination was introduced into the national immunization program in Malawi. Nevertheless, rotavirus remains a major cause of diarrhea. Over the past decade, we estimate that there has been a 36% reduction in the incidence of RVGE among children under 5 years old presenting to QECH, which is driven by the reduction in incidence in <1-year-olds. While the incidence of RVGE cases among infants decreased by over 50%, incidence has increased slightly among 1-year-olds and has more than tripled among 2-<5-year-olds, although the latter age group still represents less than 10% of all RVGE cases (range: 2-10 cases per year). Vaccine coverage has remained high, with over 95% of infants receiving at least one dose of the RV1 vaccine and over 90% receiving both doses, according to data from test-negative controls. There was a slight decrease in vaccine coverage during the COVID-19 pandemic, particularly for the second dose, which fell to approximately 80% in October 2020. We also noted substantial declines in healthcare seeking for diarrhea before, during, and after the COVID-19 pandemic, as indicated by the number of rotavirus-negative AGE cases presenting to QECH, highlighting the importance of controlling for trends in surveillance when estimating the impact of rotavirus vaccine introduction.

Our estimate of the overall impact of rotavirus vaccination in Malawi is similar to previous estimates from the first four years after vaccine introduction in Malawi [9,10] and for countries in the Global Rotavirus Surveillance Network [23], but lower than the median estimate reported for high-mortality countries in a recent review [24]. It is notable that overall vaccine impact has plateaued since 2016 and may have even decreased over the past few years. Vaccine coverage in our study population has remained high over this time period, suggesting that the reduced vaccine impact is primarily attributable to the lower vaccine effectiveness in Malawi [8]. Nevertheless, other studies have found that the severity of hospitalized RVGE cases has decreased since the introduction of RV1 [20], and infant mortality from all-cause diarrhea has also declined [25]. Thus, there may be additional benefits to vaccine introduction that have not been quantified in this analysis.

We observed only a slight drop in rotavirus vaccine coverage following the COVID-19 pandemic, primarily affecting coverage with the second dose. This small decrease in coverage is similar to that reported for the African region, based on the WHO and UNICEF Estimates of National Immunization Coverage (WUENIC) data [14]. However, larger reductions in healthcare-seeking for immunization services and other types of healthcare were reported for Malawi based on health service utilization data [15]. It is possible that our estimates of vaccine coverage are higher and less susceptible to pandemic-related disruptions, since they are based on data from test-negative controls who sought care at QECH, and the number of rotavirus-negative AGE cases dropped by over 50% between 2019 and 2021 (Table 1). Nevertheless, the vaccine coverage among test-negative controls should be representative of the population from which the rotavirus-positive cases are detected, which is necessary to provide unbiased estimates of vaccine effectiveness [26–28].

There are a variety of ways to control for trends in disease incidence when estimating the overall effectiveness of vaccine introduction, i.e. vaccine impact. Numerous studies have estimated rotavirus vaccine impact by comparing the proportion of AGE cases that tested positive for rotavirus before and after vaccine introduction, e.g. [29,30]. The benefit of this approach is that it is relatively insensitive to inconsistencies in surveillance effort over time (provided there is year-round surveillance to account for seasonal changes in the prevalence of different diarrheal pathogens [31]), and it allows for the inclusion of sentinel surveillance sites that may only have data from the pre- or post-vaccination period. Changes in healthcare access and utilization as well as other factors that may bias estimates of vaccine impact (e.g. improvements in hygiene and sanitation) are controlled for to some extent, since they affect both the numerator and denominator of the pre- and post-vaccination rotavirus prevalence. However, this approach has limited power to examine trends in rotavirus patterns and vaccine impact over time. Another common approach is to use interrupted time-series models estimate overall vaccine effectiveness, e.g. [9,29,32]. While this approach is able to assess trends in vaccine impact over time and as a function of vaccine coverage, it is still necessary to control for seasonality and secular trends in disease incidence over time.

Our analysis highlights the importance of using data on rotavirus-negative cases to control for surveillance trends and healthcare-seeking patterns, as surveillance effort at QECH increased following the introduction of RV1 in 2012 and has been decreasing over the past decade; failure to control for these patterns would lead to underestimation of vaccine impact in 2013-2015 and overestimation of vaccine impact in recent years. We assume that the decline in rotavirus-negative cases reflects decreased healthcare-seeking and/or surveillance effort at QECH, but it is also possible that it reflects a true decline in diarrhea incidence due to social distancing and other behavioral changes during the COVID-19 pandemic, as has been observed in other countries [33,34]. Regardless of the mechanism underlying the decline, our estimates of rotavirus vaccine impact are likely to remain unchanged, since the effect of social distancing is independent of vaccine introduction and likely to be similar across diarrheal pathogens.

In this analysis, we use a transmission dynamic model to predict the number of rotavirus cases pre- and post-vaccine introduction [12], and we compare the number of observed RVGE cases to that predicted by the model in the absence of vaccination to estimate vaccine impact. This approach is similar to an interrupted time-series model, but can account for other changes to the dynamics of rotavirus transmission, e.g. from changes in the birth rate or population demography over time [35]. Furthermore, by showing that the models incorporating vaccination can reproduce the observed post-vaccination dynamics of rotavirus [12], including for out-of-sample data not used for model fitting, we are able to test different hypotheses about the nature of vaccine-induced immunity. We have shown that such models are also able to reproduce the post-vaccination dynamics of rotavirus across other settings, including the US and Ghana [36,37]. Thus, the models provide a validated platform for predicting the potential impact of strategies to improve vaccine performance, including the introduction of next-generation rotavirus vaccines [38].

Nevertheless, our analysis has a number of limitations that warrant consideration when interpreting the results. As noted above, we assume test-negative AGE cases are representative controls for estimating vaccine coverage and healthcare seeking patterns. Furthermore, we assume that vaccination did not affect the incidence of non-rotavirus AGE, which may not be true if prevention of rotavirus infection decreases vulnerability to other enteropathogens and/or if diagnostic test sensitivity is low. By comparing post-vaccination RVGE cases to the number predicted by our transmission model, we assume that the model is accurately capturing the underlying natural history of rotavirus infection and immunity. Finally, it is difficult to conduct formal model selection for the four different models of vaccine protection, since there are differences in model structure and only Models 3 and 4 were formally fitted to post-vaccination data.

In conclusion, we show that the overall effectiveness of rotavirus vaccination in Malawi peaked in 2014 and has since plateaued. Since 2017, the observed number of RVGE cases at QECH was only approximately 15% lower than the incidence predicted in the absence of vaccination despite high vaccine coverage. The COVID-19 pandemic led to a slight decline in vaccine coverage, particularly for the second dose of RV1, but coverage has since returned to pre-pandemic levels as of mid-2021. Novel approaches are needed to improve rotavirus vaccine performance in low-income settings such as Malawi. Our previously published models of rotavirus vaccine impact in Malawi were able to reproduce the post-vaccination dynamics, particularly for the models assuming heterogeneity in vaccine response and/or waning of vaccine-induced immunity. These models provide a validated platform for assessing strategies to improve rotavirus vaccine impact in Malawi.

## Data Availability

All data produced in the present study are available upon reasonable request to the authors.

## Acknowledgements

This work was supported by funding from the US National Institutes of Health/National Institute of Allergy and Infectious Diseases (R01AI112970 to VEP), the Wellcome Trust (Programme grant number: 091909/Z/10/Z), the Bill and Melinda Gates Foundation (OPP1180423 and INV-046917), and US Centres for Disease Control and Prevention funds through the World Health Organization (2018/815189-0), and the UK National Institute for Health and Care Research (NIHR) Global Health Research Group on Gastrointestinal Infections at the University of Liverpool using UK aid from the UK Government to support global health research (NIHR133066). Nigel Cunliffe is a NIHR Senior Investigator (NIHR203756). Daniel Hungerford was funded through a NIHR Post-doctoral Fellowship (PDF-2018-11-ST2-006). Nigel Cunliffe, Khuzwayo C Jere and Daniel Hungerford are also affiliated with the NIHR Health Protection Research Unit in Gastrointestinal Infections at the University of Liverpool, a partnership with the UK Health Security Agency in collaboration with the University of Warwick. The funders had no role in study design, data collection and analysis, decision to publish, or preparation of the manuscript. The content is solely the responsibility of the authors and does not necessarily represent the official views of the National Institutes of Health, the NIHR, the Department of Health and Social Care, the UK government or the UK Health Security Agency.

---

**Table S1.** Model parameters.

| Fixed parameters | Variable | Value | Source |
| --- | --- | --- | --- |
| Birth rate | $B(t)$ | 0.0366 to 0.0550 year<sup>-1</sup> | [39] |
| Duration maternal immunity | $1/\omega_M$ | 26 weeks | [40] |
| Duration of infectiousness |  |  |  |
| First infection | $1/\gamma_1$ | 1 week | [41] |
| Subsequent infections | $1/\gamma_2$ | 0.5 week | [42,43] |
| Duration of temporary immunity | $1/\omega$ | 13 weeks | [44], assumption |
| Relative risk of reinfection |  |  |  |
| Following first infection | $\sigma_1$ | 0.62 | [45,46] |
| Following second infection | $\sigma_2$ | 0.35 | [45,46] |
| Relative infectiousness |  |  |  |
| Following first infection | $\rho_1$ | 0.5 | [45,46] |
| Following second infection | $\rho_2$ | 0.1 | [47] |
| Estimated parameters | Variable | Maximum a posteriori estimate (95% credible interval) |  |
| Basic reproductive number | $R_0 = \beta_0/\gamma_1$ | 78.8 (70.5-96.2) | [12] |
| Reporting rate (mean) | $h$ | 0.017 (0.016-0.018) | [12] |
| Amplitude of seasonality in transmission | $b$ | 0.174 (0.113-0.294) | [12] |
| Phase shift of seasonal transmission | $\phi$ | 6.9 (4.0-11.2) weeks | [12] |

**Table S2.**
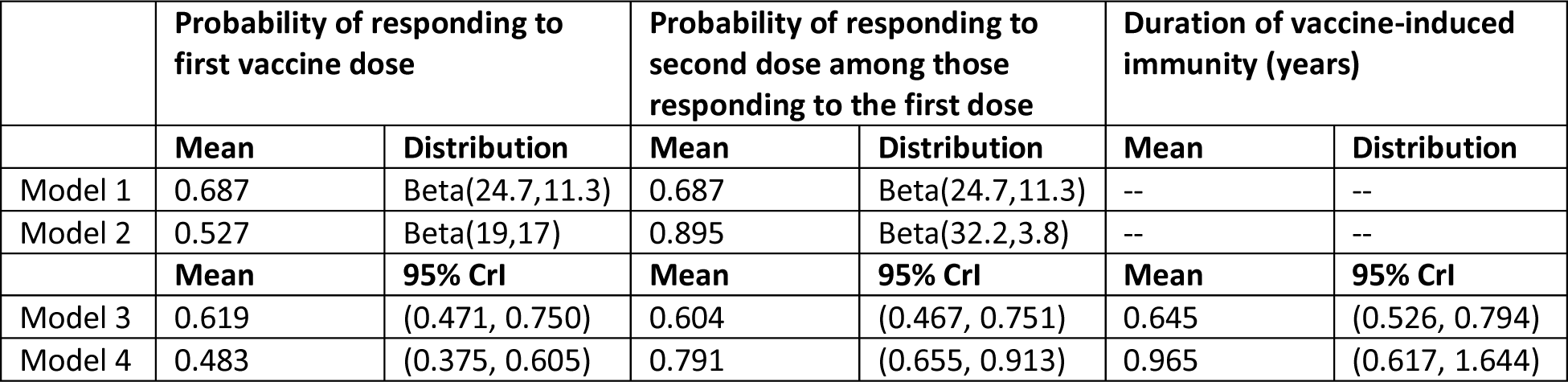
Vaccine parameters. Models 1 and 2 assume no waning of vaccine-induced immunity; the probability of responding to each vaccine dose was estimated from seroconversion data from the RV1 vaccine trial in Malawi [8]. Vaccine-related parameters for Models 3 and 4 were estimated by fitting to the post-vaccination time series of RVGE cases at Queen Elizabeth Central Hospital through August 2017 [12].

|  | Probability of responding to first vaccine dose |  | Probability of responding to second dose among those responding to the first dose |  | Duration of vaccine-induced immunity (years) |  |
| --- | --- | --- | --- | --- | --- | --- |
|  | Mean | Distribution | Mean | Distribution | Mean | Distribution |
| Model 1 | 0.687 | Beta(24.7,11.3) | 0.687 | Beta(24.7,11.3) | -- | -- |
| Model 2 | 0.527 | Beta(19,17) | 0.895 | Beta(32.2,3.8) | -- | -- |
|  | Mean | 95% CrI | Mean | 95% CrI | Mean | 95% CrI |
| Model 3 | 0.619 | (0.471, 0.750) | 0.604 | (0.467, 0.751) | 0.645 | (0.526, 0.794) |
| Model 4 | 0.483 | (0.375, 0.605) | 0.791 | (0.655, 0.913) | 0.965 | (0.617, 1.644) |

**Table S3.** Overall vaccine effectiveness estimates by year. The overall effectiveness (OE) is calculated based on the observed number of RVGE cases compared to the model-predicted incidence of RVGE with no vaccination in each year following vaccine introduction.

| Year | Age group |  |  |  |
| --- | --- | --- | --- | --- |
|  | All ages <5 years old | <1 year old | 1-<2 years old | 2-<5 years old |
| 2012* | 2.7% | 11.1% | -44.7% | -49.1% |
| 2013 | 33.3% | 48.5% | -19.5% | -320.5% |
| 2014 | 55.5% | 66.0% | 22.7% | -68.4% |
| 2015 | 26.6% | 48.1% | -37.1% | -387.3% |
| 2016 | 16.6% | 36.4% | -55.6% | -444.4% |
| 2017 | 18.8% | 34.4% | -11.5% | -1150.2% |
| 2018 | 11.2% | 37.1% | -70.7% | -741.9% |
| 2019 | 16.7% | 37.7% | -55.0% | -610.0% |
| 2020<sup>#</sup> | -15.0% | 39.9% | -214.4% | -1134.4% |
| 2021 | 17.6% | 40.9% | -55.7% | -562.2% |
| 2022<sup>§</sup> | 1.4% | 24.0% | -100.2% | 100.0% |
| **2013-2022<sup>§</sup>** | **36.0%** | **52.5%** | **-18.6%** | **-336.4%** |

\*Rotarix vaccine was introduced on October 29, 2012.
<sup>#</sup> Excludes the period from April 5 to October 10 when surveillance was halted because of the COVID-19 pandemic.
<sup>§</sup> Limited to data through June 25, 2022.

**Table S4.**
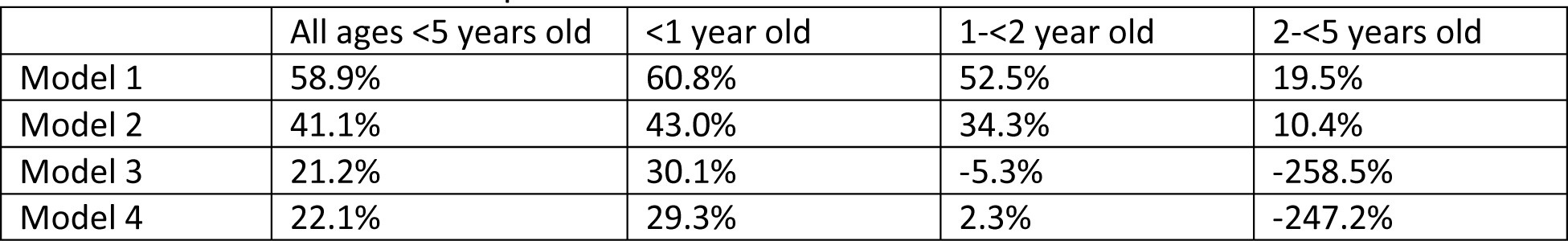
Model-predicted overall vaccine effectiveness. The overall effectiveness (OE) predicted by each of the four vaccination models compared to the model-predicted incidence of RVGE with no vaccination is presented for the best-fit models.

**Figure S1.**
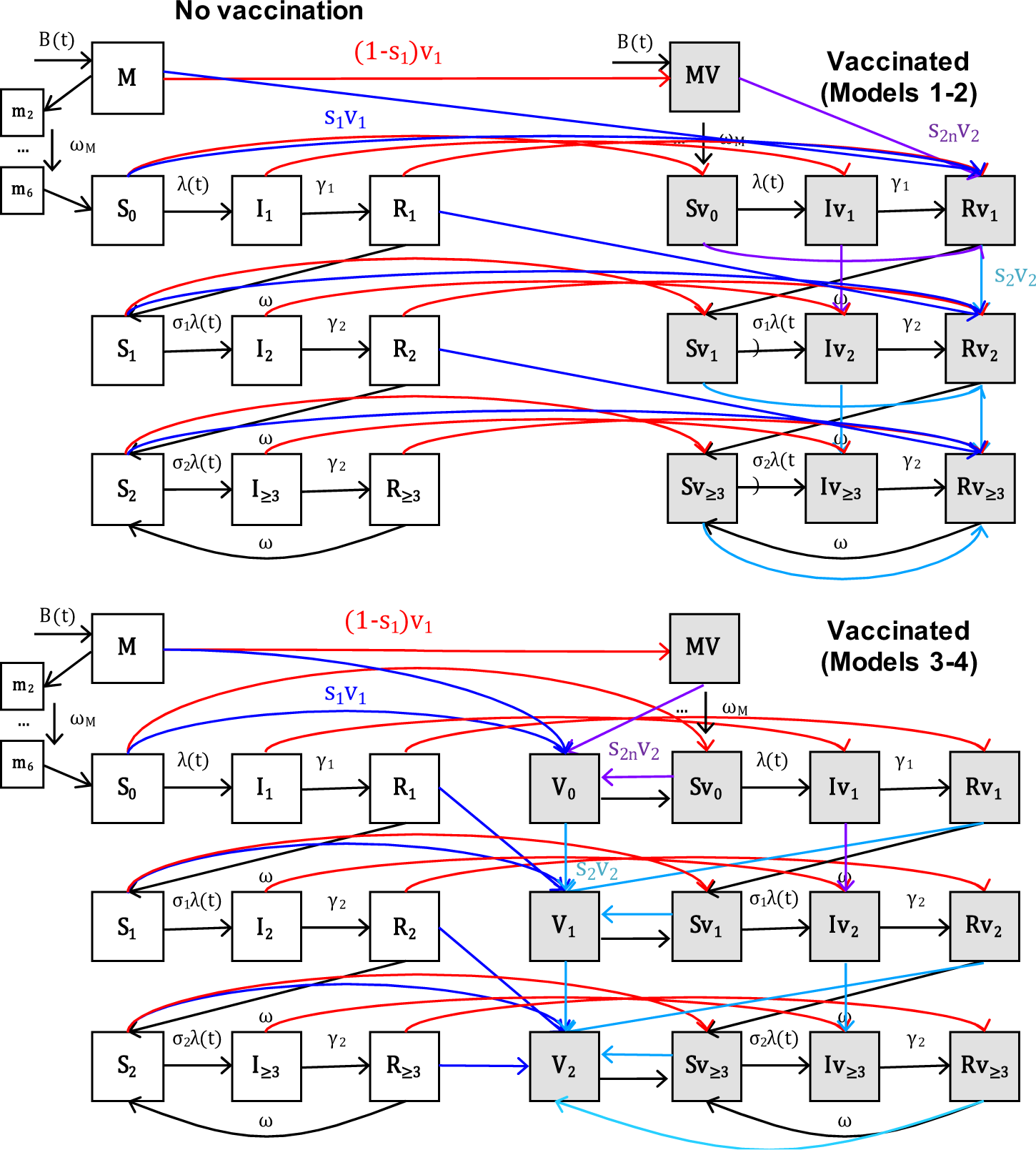
Compartmental diagram of transmission models. Boxes represent the various model states (unvaccinated in white, vaccinated in grey), while the lines represent the movements between model states for (top) Models 1 and 2 (assuming vaccine-induced immunity is comparable to immunity from natural infection) and (bottom) Models 3 and 4 (assuming waning of vaccine-induced immunity). The blue and turquoise lines represent the movement of individuals who respond to the first and subsequent doses of rotavirus vaccine, respectively, while the red lines represent the movement of individuals who fail to respond to the first dose. The purple lines represent the probability of responding to the second dose among those who failed to respond to the first dose when we assume heterogeneity in vaccine response (Models 2 and 4). Individuals who fail to respond to subsequent doses remain in their respective vaccinated compartments. Adapted from [12].

**Figure S2.**
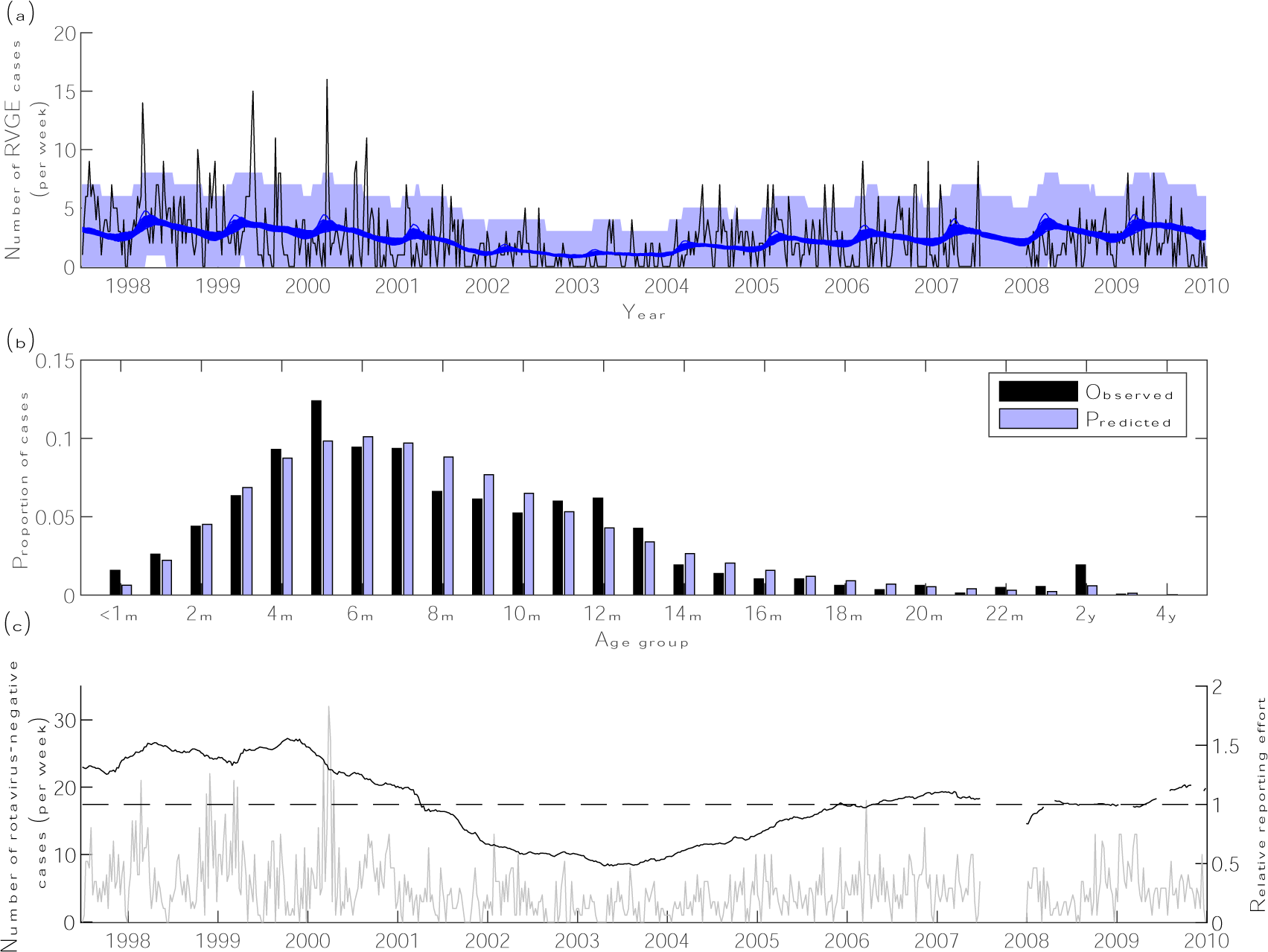
Weekly timeseries of rotavirus-positive and negative cases and fitted model for pre-vaccination period, July 1997-December 2009. (a) The number of rotavirus-associated gastroenteritis (RVGE) cases per week diagnosed at Queen Elizabeth Central Hospital in Blantyre, Malawi is plotted in black, while the fitted models are plotted in blue for 100 samples from the posterior distribution of model parameters. The blue shaded region represents the 95% prediction intervals assuming the observed number of cases per week are Poisson distributed with a mean equal to the model-predicted weekly average number of cases. (b) The age distribution of observed RVGE cases (black bars) is plotted alongside the model-predicted age distribution of RVGE cases (blue bars). (c) The number of rotavirus-negative acute gastroenteritis cases per week are plotted in grey on the left axis, while the relative reporting effort over time is plotted in black on the right axis; the black dashed line shows a relative reporting effort of 1. The reporting effort is calculated from the 105-week (two-year) moving average of the number of rotavirus-negative cases divided by the average for the entire time period.

**Figure S3.**
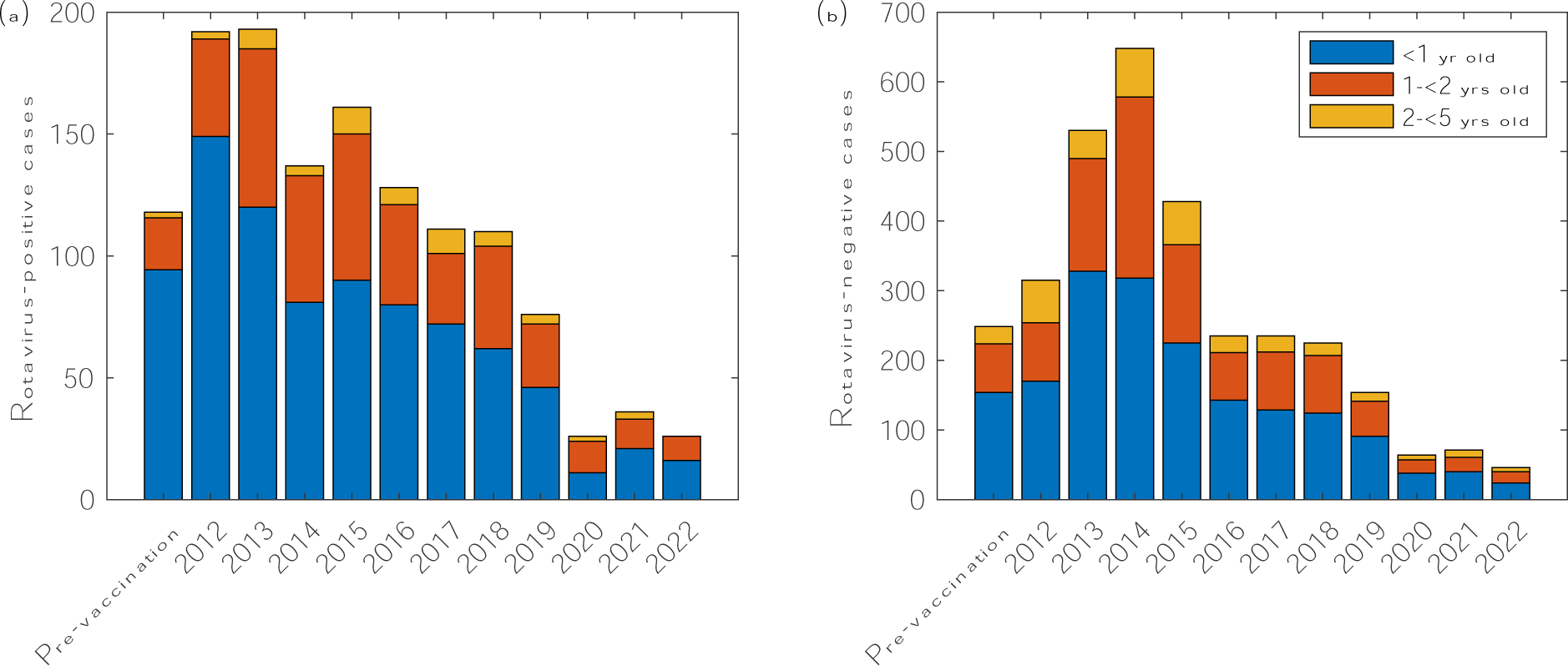
Annual cases of rotavirus-positive and rotavirus-negative acute gastroenteritis by age group, 1997-2022. The number of (a) rotavirus-positive and (b) rotavirus-negative acute gastroenteritis cases presenting to Queen Elizabeth Central Hospital in Blantyre, Malawi are plotted by year and age group (blue <1 year old; red 1-<2 years old; yellow 2-<5 years old). The first bar represents the average annual number of cases from the pre-vaccination period (June 1997-December 2009); data from 1997 and 2007 were excluded because surveillance was only conducted for six months in these years.

**Figure S4.**
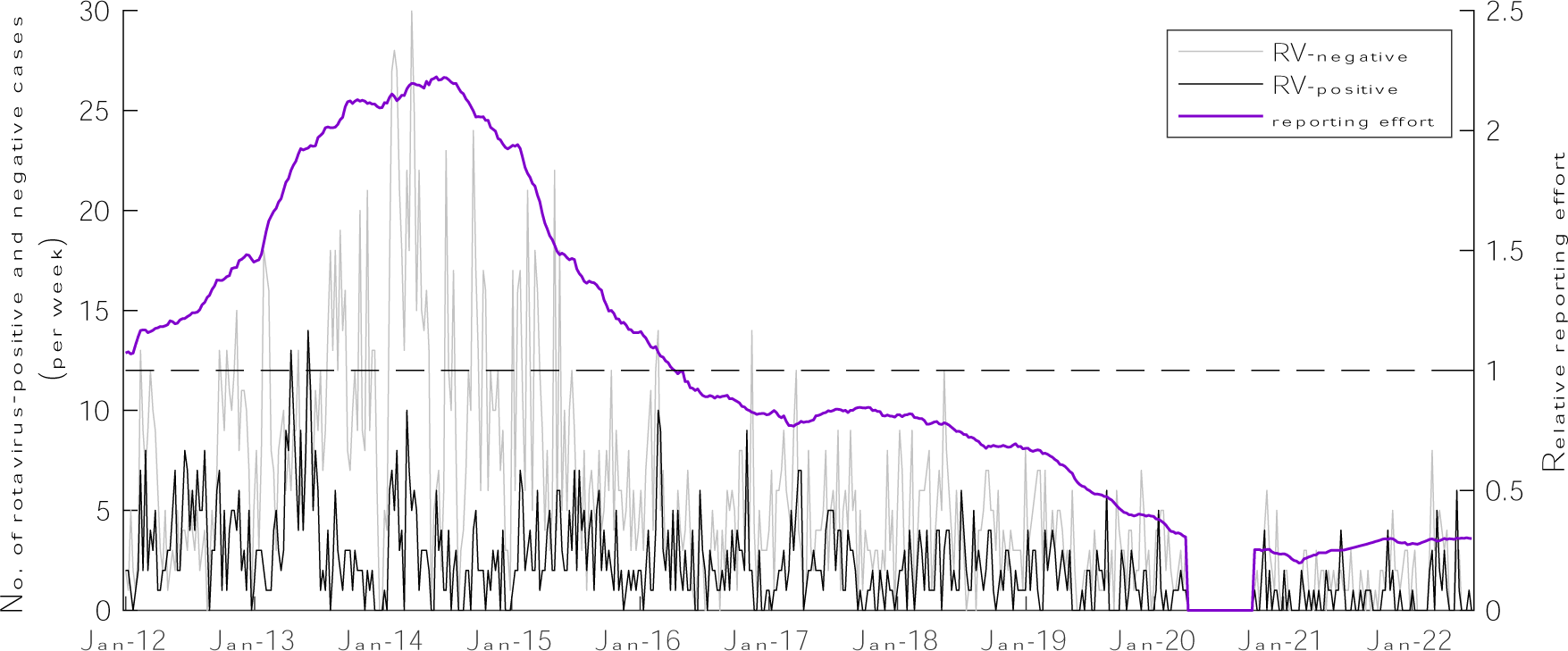
Weekly timeseries of rotavirus-positive and negative cases and relative reporting effort, January 2012-June 2022. The number of rotavirus-positive (black) and rotavirus-negative (grey) acute gastroenteritis cases per week diagnosed at Queen Elizabeth Central Hospital in Blantyre, Malawi are plotted on the left axis. The relative reporting effort over time (purple) is calculated from the 105-week (two-year) moving average of the number of rotavirus-negative cases divided by the average for the entire time period and is plotted on the right axis. The black dashed line shows a relative reporting effort of 1.

**Figure S5.**
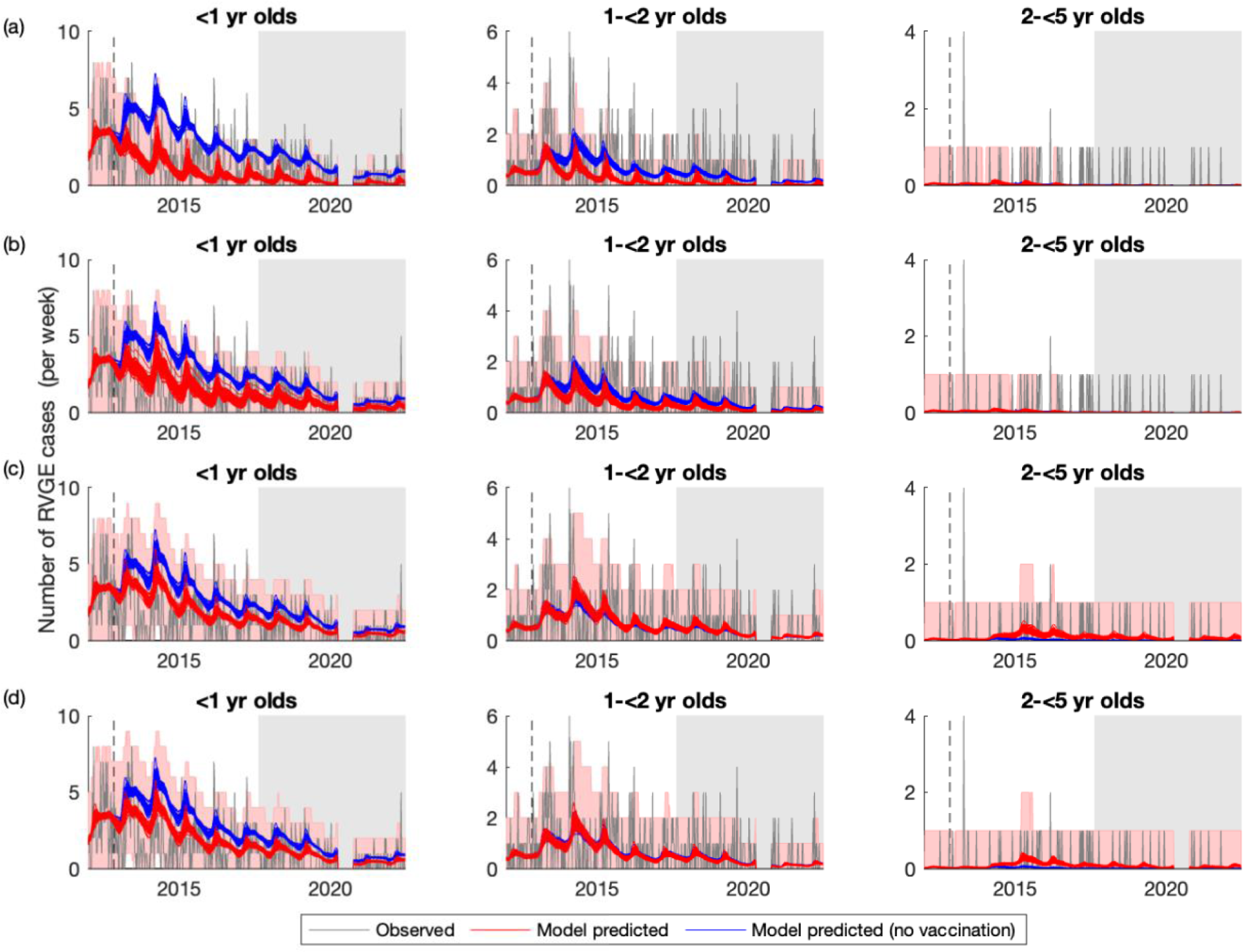
Weekly timeseries of observed and model-predicted rotavirus gastroenteritis cases by age group at Queen Elizabeth Central Hospital, January 2012-June 2022. The observed number of RVGE cases per week for three different age groups (<1 year old, left; 1-<2 year old, middle; 2-<5 year old, right) is plotted in grey, while model predictions for the average weekly number of RVGE cases given current estimates of vaccine coverage (red lines) and assuming no vaccination (blue lines) are plotted for 100 samples from the posterior distribution of model parameters for (a) Model 1, (b) Model 2, (c) Model 3, and (d) Model 4. The red shaded region represents the 95% prediction intervals assuming the observed number of cases per week are Poisson distributed. The dashed vertical line shows the week of vaccine introduction, while the light grey shaded region shows the out-of-sample validation period.

## Notes

### Competing Interest Statement

The authors have declared no competing interest.

